# Evaluating Chinese Handwriting Performance of Primary School Students Using the Smart Handwriting Analysis and Recognition Platform (SHARP)

**DOI:** 10.1101/2022.02.19.22270984

**Authors:** Cecilia W. P. Li-Tsang, Tim M. H. Li, C.N. Yang, Howard W. H. Leung, Eve W. L. Zhang

**Affiliations:** Department of Rehabilitation Sciences, The Hong Kong Polytechnic University, Hung Hom, Kowloon, Hong Kong (SAR), China; Department of Computer Science, City University of Hong Kong, Kowloon Tong, Kowloon, Hong Kong (SAR), China; Department of Psychiatry, Faculty of Medicine, The Chinese University of Hong Kong

**Author notes:** Corresponding author: Adjunct Professor Cecilia Li-Tsang W.P., Department of Rehabilitation Science, The Hong Kong Polytechnic University, Hung Hom, Kowloon, Hong Kong., /.

**Keywords:** Handwriting performance, Neurodevelopmental problems, Computer-based handwriting assessment

## Abstract

**Background:** Handwriting is one of the fundamental skills for school-aged children, and it is an intricate and interacting process. An innovative computerized handwriting assessment system, the Smart Handwriting Analysis and Recognition Platform (SHARP) was used to provide a comprehensive and quantitative analysis of Chinese handwriting performance and identify children with handwriting difficulties objectively, accurately, and efficiently.

**Aims/Objectives:** The purpose of this study is to provide quantitative data on children’s Chinese handwriting process and products using SHARP; to investigate the handwriting difficulties in children with neurodevelopmental problems, and to explore the associations between handwriting and sensorimotor performance, including fine motor skills, visual perceptual skills, visual-motor integration, and oculomotor proficiency.

**Material and Methods:** Typically developing students and students with neurodevelopmental problems were recruited through teachers and therapists from six primary schools in Hong Kong. SHARP and tests of sensorimotor skills were performed.

**Results:** The quantitative analysis using the SHARP system showed a progressive developmental change in handwriting performance among the typically developing children at their primary education. There were also significant differences between the typically developing students group and those with neurodevelopmental problems, both in handwriting speed and writing errors. Significant associations between the handwriting process parameters and fine motor skills were identified, while handwriting product parameters were closely correlated with visual-perceptual skills of these typical developing students.

**Conclusions and Significance:** SHARP appears to provide a comprehensive, quantitative and objective evaluation of students’ handwriting performances, thus assisting both teachers and parents in comparing students’ performance objectively, thus identifying students with handwriting difficulties as early as P1 level. It was observed that handwriting performance has strong relationships with young children’s visual perceptual skills and fine motor skills. The findings could help to facilitate early intervention for those students with handwriting problems.

## Introduction

Handwriting is one of the fundamental skills for school-aged children. Handwriting is an intricate and interacting process, which was shown to associate with sensorimotor abilities such as visual perception, visual-motor integration, in-hand manipulation, and oculomotor abilities [1]. Deficits in these abilities could affect children’s handwriting speed, writing stability, and letter quality [2]. For example, visual perceptual skills are necessary for children to write inside the grids correctly [2]. Visual-motor integration (VMI) is considered a prerequisite skill before learning handwriting and a predictor of handwriting quality [3]. Oculomotor control is another essential eye movement ability, a fundamental skill to plan and guide hand movement [1, 4]. It is crucial to investigate the associations between handwriting performance and the underlying sensorimotor skills.

Around 5-33% of children have handwriting problems [5]. The problem is more common in children with neurodevelopmental problems such as autism spectrum disorder (ASD), attention deficit hyperactivity disorder (ADHD), and specific learning difficulties (SpLD), as they have more sensorimotor challenges [6]. Assessment of sensorimotor functions is well developed and validated, whereas handwriting assessment has been evolving over the past decades. The handwriting performance of school-aged children was simply assessed using pencil-and-paper near-point copying tasks [7]. Falk, Tam [8] hence developed a computerized Minnesota Handwriting Assessment (MHA) to measure English handwriting speed and legibility. Li-Tsang, Wong [9] also established an evaluation tool, the Chinese Handwriting Analysis System (CHAS), to assess Chinese handwriting speed and accuracy. Previous computerized handwriting analysis systems adopted traditional approaches that rely on predefined algorithms to analyze handwriting products [9]. These traditional computerized approaches might neglect some subtle non-intuitive variables that contribute to a more accurate model in handwriting evaluation. Thus, Li and colleagues have further established the Smart Handwriting Analysis and Recognition Platform (SHARP), which utilizes deep learning techniques in analyzing the handwriting product [10]. The deep learning approach has been regarded as superior to traditional algorithms, with higher efficiency in development, better accuracy, and supreme flexibility [11].

Handwriting assessments often take a product-oriented approach [12] and adopt speed and accuracy as main constructs [9]. CHAS and SHARP have been developed to depict the readability of Chinese characters, including wrong stroke, additional stroke, missing stroke, concatenated stroke, reverse stroke, and wrong stroke sequence as key parameters [9, 10]. Chinese is a logographic language, which is composed of different characters. Each character is formulated by one or several radicals. Each radical is composed of multiple strokes of various kinds. The accuracy of the strokes arrangement is essential for the accuracy of the character. Even a slight difference in stroke arrangements could represent different meanings; for example, the word of sky would become husband when a stroke is extended to cross the upper horizontal stroke. In Chinese handwriting, the location, the proportion, the direction, and the size of strokes are all vital to correct writing.

This study aims to (1) provide quantitative data on children’s Chinese handwriting process and products using SHARP, (2) investigate the handwriting difficulties in children with neurodevelopmental problems, and (3) explore the associations between handwriting and sensorimotor performance, including fine motor skills, visual perceptual skills, visual-motor integration, and oculomotor proficiency.

## Materials and Methods

### Participants

Cross-sectional research design was adopted, and a stratified sample was collected based on the geographical regions of the city and population. A total of six primary schools were randomly selected from four major geographic regions according to their proportionality to the total population. A class from each grade (from grades 1-6) was randomly selected from the sampled primary schools. All students were included in the study except students with (a) any physical disabilities affecting the upper limb function, (b) any vision, hearing, and speech disorders such as visual impairment, deaf and hard of hearing, and communication disorder, and (c) who are not native Chinese.

A total of 507 students (mean age = 9.32, SD = 1.87, 49% male) were recruited for the study. Upon their parents’ consent and ethical approval from the University, they were invited to participate in the handwriting assessment. Among the subjects, 212 students (mean age =8.44, SD=7.6, 49% male) were consented to participate in the sensorimotor assessments for their visual perception skills, oculomotor control, fine motor skills, and visual-motor integration. Students with neurodevelopmental problems were recruited through teachers and therapists either from the six schools or from other sources. A total of eighty-four students with neurodevelopmental problems (mean age = 9.57, SD = 1.73, 76% male) were recruited.

All participants were informed about their rights to participate in this study, and written consent was obtained from their parents before the study. The study was approved by the Human Subjects Ethics Sub-committee of The Hong Kong Polytechnic University.

### Measures

#### Chinese Handwriting Assessment

The handwriting assessment was conducted in a classroom setting during school days. Students were given a digital tablet and a paper template. They were asked to follow the instruction from the examiner, turn to the template page, and start copying. A student had to copy 90 Chinese characters from left to right and from up to down as fast and accurately as possible but was reminded to write clearly within the grid lines.

The “SHARP” system was installed into a portable A4-sized tablet for data collection and recording [10]. The handwriting data used in this research was recorded with handwriting tablets Digit Note (Type PH-1410). The data collected for each subject were uploaded to the cloud platform and monitored by a team of software engineers and health professionals. MyScript handwriting recognition technology was adopted for Chinese character recognition to provide the number of identified words [13]. Information on the handwriting process, including on-paper time (ground time), on-air time (air time), air/ground time ratio, writing speed, SD of the writing time per character, pen-tip pressure, and SD of pressure were recorded. It could also detect the number of words out of the grid, word size, and SD of size.

A total of 90 individual Chinese characters were selected from “A Study of the Chinese Characters Recommended for the subject of the Chinese language in Primary Schools” [14]. The selection of characters was based on the frequency level, the character’s structure, and common strokes. The characters were selected among the most commonly used characters by primary school students. All the common strokes listed in “Path to Mastery of Chinese Characters: Chinese Character Writing Courseware” [15] were present in the 90 characters. Twenty characters were selected to represent handwriting accuracy.

#### Tests of Sensorimotor Skills

Four sensorimotor performance components included visual-motor integration, visual perception, fine motor skills, and ocular motor skill. The Beery-Buktenica Developmental Test of Visual-Motor Integration-6th Edition (Beery VMI) was used for examining visual perception and motor abilities coordination [16]. The VMI consists of 30 geometric forms, and the students were instructed to copy a series of geometric designs with increasing complexity. Their drawings were scored based on standard criteria; a higher score indicated a greater function in visual-motor integration. The test has high content and person reliability from the Rasch-Wright results, with a total group item separation of 1.00 and a total group person separation of .96. The interscorer reliability of VMI is .93. The concurrent, predictive, and content validity were established for previous versions.

Test of Visual-Perceptual Skills (non-motor), Third Edition (TVPS-3) was used for visual perception skills testing [17]. There are seven subtests, including Visual Discrimination (DIS), Visual Memory (MEM), Visual-Spatial Relationships (SPA), Form Constancy (CON), Visual Sequential Memory (SEQ), Visual Figure-Ground (FGR), Visual Closure (CLO). The students were instructed to view a picture with multiple choices and select the correct answer. The total score was the sum of seven subtests ranging from 0 to 112; a higher score indicates a greater function in visual perception. TVPS-3 showed to have satisfactory test-retest reliability (ICC = .92) and good internal consistency (α = .71- .89) [18].

The Bruininks-Oseretsky Test of Motor Proficiency-2nd Edition (BOT-2) [19] was used to measure fine motor skills. Among eight subtests, three subtests Fine Motor Precision (FMP), Fine Motor Integration (FMI), and Manual Dexterity (MD), were chosen to be used for fine motor function testing. The tests have high internal consistency (α = .78– .97), test-retest reliability (r = .52– .95), and inter-rater reliability (κ = .92) [19].

The brief developmental eye movement test (DEM) [20] was used to evaluate the ocular-motor control and rapid automatized naming (RAN) of the students. It consists of two subtests, vertical subtest (DEM-V) and horizontal subtest (DEM-H). The students were instructed to read the digital number displayed in a vertical array in the vertical test. While in a horizontal subtest, the students were requested to read the digital number displayed in a horizontal array with uneven distribution without the assistance of a finger. The performance was measured in time to the nearest 0.01 second. The ratio (DEM-R) was calculated. A person with a high ratio and high score in the horizontal subtest was classified as having ocular motor dysfunction. Normal ratio and high scores in horizontal and vertical subtests imply that a person had difficulties with the automaticity in number calling skills. With a high ratio and high scores in both subtests, a person had difficulties in both ocular motor skill and automaticity. The DEM demonstrated a good to excellent reliability (ICC > .75), good consistency with diagnosis (k = .56 - .77) [21].

### Statistical Analysis

All statistical computations were performed by statistical software R (R for Windows, V.3.4.3). Descriptive statistics for continuous variables were illustrated by means and SD; categorical variables were represented by numbers and percentages. Handwriting performance was compared among the typically developing students from grades 1-6 using a one-way analysis of variance. The handwriting characteristics (independent variables) of typically developing students and students with neurodevelopmental problems (dependent variable) were compared by binary logistic regression. The odds ratios and 95% confidence intervals were presented. Students’ gender and grade were included as independent variables for confounder adjustment. Linear regression was used to examine the associations between sensorimotor performance (including visual-motor integration, visual-perceptual skills, fine motor, and eye movement development) and handwriting scores (i.e., handwriting process and handwriting product).

## Results

### Handwriting Performance of Typically Developing Students

Table 1 shows the handwriting process and product of typically developing students from primary grade 1 to 6 based on the results generated by SHARP. The handwriting process, including ground time, air time, air/ground ratio, speed, SD of writing time per character, pen pressure, SD of pressure, had a significant progression of improvement from grades 1-6. For the performance on the handwriting products, senior students had a reduced number of writing out of the grid, wrong strokes, additional strokes, reverse strokes, and wrong stroke sequence, and increased identified words when compared to junior students. The size of words was smaller in senior students when compared to junior students. The junior students had more air time than the ground time during the writing process, while the senior students showed similar air and ground time. There was no significant improvement in missing stroke and concatenated stroke from grades 1-6.

**TABLE 1.** Characteristics and handwriting performance of typically developing participants.

|  | P.1 | P.2 | P.3 | P.4 | P.5 | P.6 | *p* |
| --- | --- | --- | --- | --- | --- | --- | --- |
| Gender | n (%) | n (%) | n (%) | n (%) | n (%) | n (%) |  |
| Male | 56 (22) | 47 (19) | 36 (14) | 42 (17) | 35 (14) | 33 (13) | .10 |
| Female | 58 (22) | 47 (18) | 40 (16) | 34 (13) | 41 (16) | 38 (15) | .14 |

|  | Mean<br>(SD) | Mean<br>(SD) | Mean<br>(SD) | Mean<br>(SD) | Mean<br>(SD) | Mean<br>(SD) |  |
| --- | --- | --- | --- | --- | --- | --- | --- |
| Age | 6.99<br>(0.44) | 8.05<br>(0.53) | 9.13<br>(0.51) | 10.13<br>(0.58) | 11.15<br>(0.69) | 12.11<br>(0.57) | <.001*** |
| Handwriting process |  |  |  |  |  |  |  |
| Ground time (min) | 5.35<br>(1.55) | 4.92<br>(1.78) | 3.19<br>(0.86) | 2.94<br>(0.89) | 2.45<br>(0.64) | 2.19<br>(0.48) | <.001*** |
| Air time (min) | 9.06<br>(3.05) | 6.89<br>(3.12) | 4.58<br>(1.40) | 4.09<br>(1.35) | 3.38<br>(1.02) | 2.94<br>(0.86) | <.001*** |
| Air/Ground time ratio | 1.77<br>(0.59) | 1.48<br>(0.51) | 1.48<br>(0.43) | 1.44<br>(0.41) | 1.43<br>(0.45) | 1.36<br>(0.38) | <.001*** |
| Speed (words/min) | 6.56<br>(1.67) | 8.22<br>(2.28) | 12.21<br>(2.79) | 13.64<br>(3.40) | 16.22<br>(3.92) | 18.32<br>(3.88) | <.001*** |
| SD of writing time per character | 5.79<br>(2.04) | 4.71<br>(2.44) | 2.80<br>(1.05) | 2.30<br>(0.88) | 1.84<br>(0.61) | 1.47<br>(0.47) | <.001*** |
| Pen pressure | 1598.31<br>(230.95) | 1652.54<br>(215.03) | 1663.61<br>(196.44) | 1560.13<br>(264.73) | 1637.90<br>(233.86) | 1534.77<br>(289.45) | .003** |
| SD of pressure | 511.00<br>(66.03) | 497.06<br>(62.87) | 500.17<br>(56.73) | 493.62<br>(56.18) | 476.96<br>(65.62) | 483.91<br>(65.11) | .006** |
| Handwriting product |  |  |  |  |  |  |  |
| Out of grid (no. of words) | 32.21<br>(19.18) | 35.84<br>(21.77) | 25.13<br>(18.42) | 24.38<br>(19.43) | 19.14<br>(18.38) | 16.46<br>(18.06) | <.001*** |
| Size (mm<sup>2</sup>) | 86.52<br>(33.62) | 74.97<br>(24.36) | 61.40<br>(22.82) | 58.27<br>(24.54) | 52.06<br>(26.25) | 37.47<br>(15.71) | <.001*** |
| SD of size (mm<sup>2</sup>) | 61.66<br>(204.39) | 62.03<br>(158.33) | 35.47<br>(41.40) | 35.52<br>(51.44) | 35.33<br>(111.84) | 18.29<br>(7.09) | .17 |
| Identified words (%) | 94.12<br>(14.73) | 92.29<br>(10.41) | 97.63<br>(3.41) | 96.91<br>(4.00) | 97.76<br>(4.50) | 98.31<br>(2.92) | <.001*** |
| Wrong stroke (%) | 33.39<br>(16.23) | 35.96<br>(18.57) | 25.12<br>(14.43) | 24.89<br>(15.69) | 19.78<br>(12.42) | 23.96<br>(15.09) | <.001*** |
| Additional stroke (%) | 42.13<br>(17.88) | 38.54<br>(17.42) | 25.93<br>(13.41) | 22.91<br>(15.86) | 17.31<br>(11.07) | 16.08<br>(12.15) | <.001*** |
| Missing stroke (%) | 13.98<br>(10.87) | 15.53<br>(12.93) | 13.79<br>(11.94) | 15.91<br>(11.73) | 13.85<br>(10.32) | 18.59<br>(14.20) | .10 |
| Concatenated stroke (%) | 26.26<br>(16.14) | 28.56<br>(18.06) | 26.22<br>(18.96) | 31.39<br>(19.79) | 27.66<br>(19.43) | 32.31<br>(20.49) | .17 |
| Reverse stroke (%) | 3.80 | 4.54 | 2.02 | 3.31 | 1.93 | 1.78 | .002** |
|  | (4.73) | (5.55) | (3.80) | (9.64) | (3.49) | (2.96) |  |
| Wrong stroke sequence (%) | 22.68<br>(11.67) | 21.18<br>(10.89) | 20.99<br>(9.06) | 22.19<br>(11.38) | 18.21<br>(9.86) | 17.61<br>(10.17) | .008** |

P = primary; SD = standard deviation; \* $p < .05$ ; \*\* $p < .01$ ; \*\*\* $p < .001$

### Comparison Between Typically Developing Students and Students with Neurodevelopmental Problems

There were significant differences between typically developing students and students with neurodevelopmental problems regarding handwriting process and product (Table 2). In the handwriting process, students with neurodevelopment problems had larger ground time, air time, SD of writing time per character, and slower writing speed than the typically developing students. For the handwriting product, students with neurodevelopmental problems had more words writing out of the grid, larger word size, less identified words, as well as more numbers of the wrong strokes, missing strokes, concatenated strokes, and reverse strokes.

**TABLE 2.**
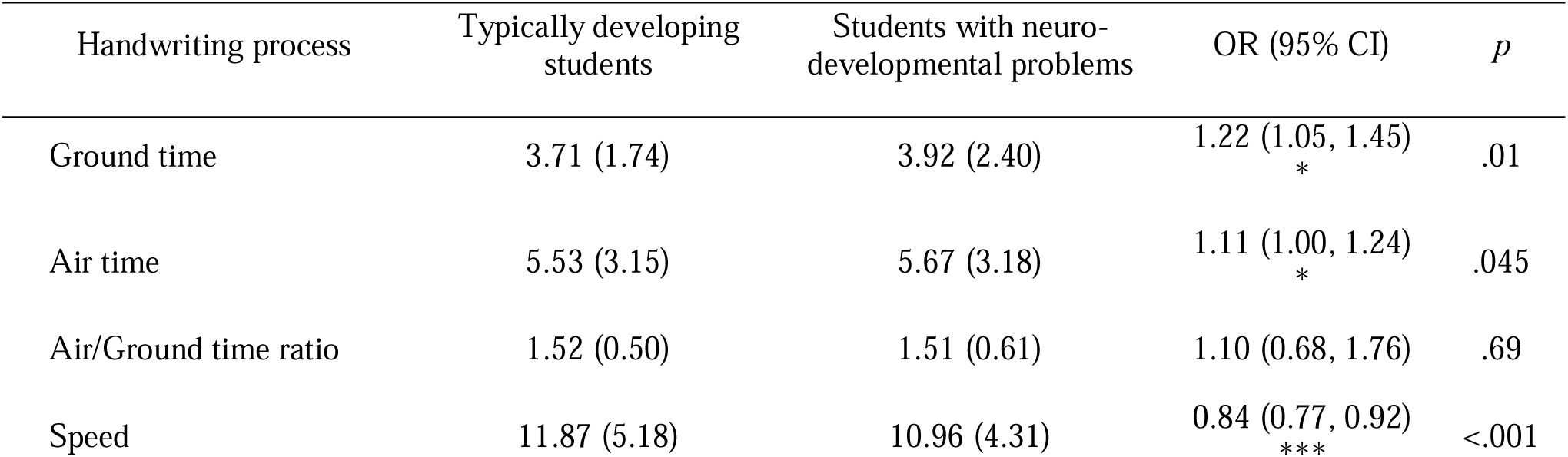
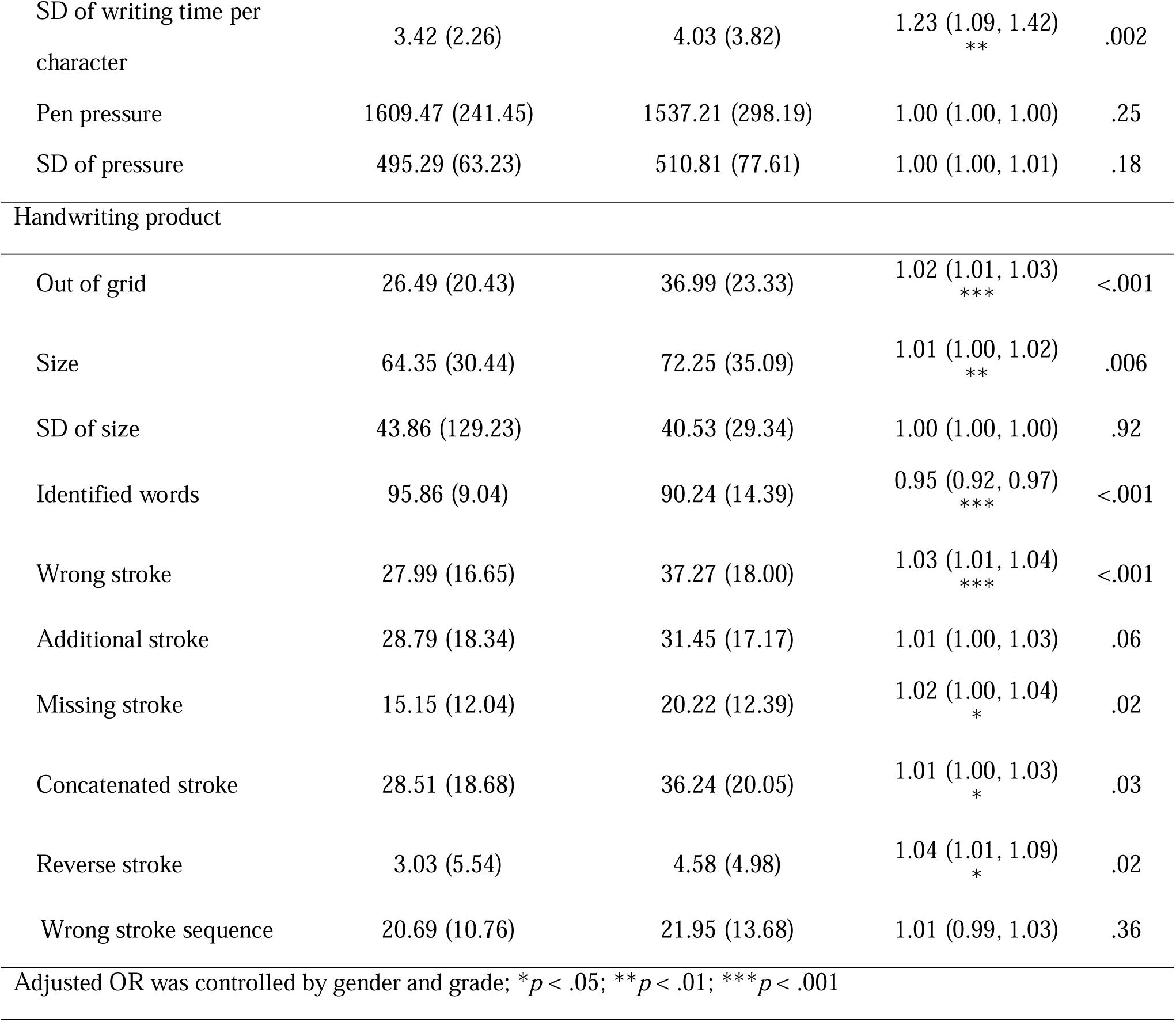
Comparison of handwriting performance between the typically developing students and students with neurodevelopmental problems.

### Relationships Between Sensorimotor Skills and Handwriting Performance

The associations between the handwriting process, handwriting product, and sensorimotor performance components are shown in Table 3. Handwriting process was found to be associated with Manual Dexterity (β = 0.08, p < 0.001) and DEM vertical time (β = -0.03, p < 0.001). Handwriting product associated with Visual Motor Integration (VMI, β = 0.09, p = 0.03), Visual Discrimination (DIS, β = 0.12, p = .0008), Visual Memory (MEM, β = 0.12, p = 0.008), Form Constancy (CON, β = 0.08, p = 0.04), DEM horizontal time (β = -0.02, p < .001). There were no significant associations between handwriting scores, Visual-Spatial Relationships (SPA), Visual Sequential Memory (SEQ), Visual Figure-Ground (FGR), Visual Closure (CLO), Fine Motor Precision (FMP), and Fine Motor Integration (FMI).

**TABLE 3.** Association between handwriting and sensorimotor performance using linear regression.

| Sensorimotor component | Handwriting process |  | Handwriting product |  |
| --- | --- | --- | --- | --- |
| | $\beta$ (95% CI) | $p$ | $\beta$ (95% CI) | $p$ |
| VMI | 0.01 (-0.05, 0.07) | 0.71 | 0.09 (0.01, 0.17) * | 0.03 |
| TVPS-4 |  |  |  |  |
| DIS | 0.04 (-0.02, 0.10) | 0.21 | 0.12 (0.03, 0.20) ** | 0.008 |
| MEM | 0.05 (-0.02, 0.11) | 0.14 | 0.12 (0.03, 0.21) ** | 0.008 |
| SPA | -0.02 (-0.07, 0.03) | 0.44 | 0.03 (-0.04, 0.09) | 0.44 |
| CON | 0.04 (-0.01, 0.10) | 0.14 | 0.08 (0.00, 0.16) * | 0.04 |
| SEQ | 0.05 (-0.02, 0.11) | 0.14 | 0.03 (-0.05, 0.12) | 0.45 |
| FGR | -0.02 (-0.07, 0.03) | 0.36 | 0.05 (-0.02, 0.13) | 0.13 |
| CLO | 0.01 (-0.04, 0.06) | 0.59 | 0.06 (-0.01, 0.13) | 0.10 |
| BOT |  |  |  |  |
| FMP | -0.02 (-0.05, 0.01) | 0.27 | 0.04 (-0.01, 0.08) | 0.09 |
| FMI | 0.02 (-0.01, 0.05) | 0.25 | 0.03 (-0.02, 0.08) | 0.22 |
| MD | 0.08 (0.03, 0.12) *** | <.001 | 0.04 (-0.02, 0.10) | 0.15 |
| DEM |  |  |  |  |
| Vertical time | -0.03 (-0.04, -0.02) *** | <.001 | -0.02 (-0.03, 0.00) | 0.07 |
| Horizontal time | -0.01 (-0.02, 0.00) | 0.05 | -0.02 (-0.03, -0.01) *** | <.001 |
| Score | -0.03 (-0.20, 0.14) | 0.74 | 0.16 (-0.07, 0.39) | 0.18 |

VMI = BEERY Visual-Motor Integration; TVPS-4 = Test of Visual-Perceptual Skills-4<sup>th</sup> Edition; DIS = Visual Discrimination; MEM = Visual Memory; SPA = Spatial Relationships; CON = Form Constancy; SEQ = Sequential Memory; FGR = Visual Figure-Ground; CLO = Visual Closure; BOT = Bruininks - Oseretsky Test; FMP = Fine Motor Precision; FMI = Fine Motor Integration; MD = Manual Dexterity; DEM = Developmental Eye Movement test; Adjusted coefficient was controlled by gender and grade; \* $p < .05$ ;

## Discussion

This study provides a quantitative handwriting profile for students in primary education using SHARP. The findings show a gradual improvement in the handwriting process parameters for students from grades 1-6. Children’s reading performance, visual processing speed, and finger dexterity skills to grasp and manipulate objects become gradually mature when they get to higher grades in school. The development of control over the large and small muscles, visual perception, fine motor skills, and in-hand manipulation skills also facilitate senior students to perform much better on handwriting speed and accuracy when compared to junior students.

Since letter copying requires the coordination of visual and motor skills in real-time, Fears and Lockman [22] suggested that younger children need more time to visually process a letter or symbol and initiate a writing action when compared with older children. This could be the reason why, in this study, the junior students spent more air time than ground time while writing. For the senior students, the difference between ground time and air time became smaller. According to Rosenblum, Weiss [23], children with poor handwriting tend to have more air time movements above the writing surface. They need more time to visually process a character or symbol and transfer it into writing compared to senior students. Therefore, the significant difference between ground time and air time may imply the occurrence of handwriting difficulty in children.

SHARP appeared to discriminate students with neurodevelopmental problems on their handwriting and discovered that they required longer time to complete the handwriting, both in ground time, air time, and SD of writing time per character, as well as slower writing speed. In the handwriting product, students with neurodevelopmental problems had more handwriting mistakes, namely,, writing out of the grid, larger word size, fewer identified words, as well as more numbers of the wrong stroke, missing stroke, concatenated stroke, and reverse stroke when compared to the typical students. Similar characteristics were observed in previous studies where participants with ADHD-LD had increased air time, slower writing speed, and higher writing speed variation when compared with their matched controls in Chinese handwriting[6].

Another advantage of SHARP is that it will automatically update the database such that it could become more sensitive and accurate to detect students with handwriting problems. Suppose the SHARP database is enhanced in the future; in that case, it will identify the patterns of handwriting errors among those students with specific neurodevelopmental disorders such as ASD, ADHD, or SpLD, and ultimately provide norms of handwriting performance for different special groups of children.

Visual-motor integration (VMI), form constancy (CON), visual discrimination (DIS), visual memory (MEM), and DEM at a horizontal time were significantly related to the handwriting product that involves legibility and accuracy. Legibility is influenced by the shape, the size, the arrangement, and the amount of space between the letters in English alphabets [24]. In Chinese characters, the parameters of the handwriting product are measured by the number of words written out of the grid, word size, and the numbers of the wrong stroke, additional stroke, missing stroke, concatenated stroke, and reverse stroke. According to Meng, Wydell [25], writing Chinese characters requires children to discriminate against the difference in the structures of characters through the visual-motor abilities to coordinate finger motion and basic visual input. This further explains why VMI and MEM were significantly related to the Chinese handwriting product. Senior students at grade 6 had a reduced number of writing out of the grid, wrong stroke, additional stroke, reverse stroke, and wrong stroke sequence, and increased identified words compared to lower grade-level students.

SHARP could provide a comprehensive and quantitative analysis of students’ handwriting processes and products. The progression of the handwriting ability in writing Chinese characters was plotted, and abnormality from normal developmental courses could be easily identified. This can also relieve the anxiety of teachers or parents regarding their children’s handwriting performance as some may not reach the developmental stage of progression yet. With accurate and early identification of students with handwriting problems using SHARP in the school setting, the teachers could then refer the student for further assessments by psychologists or occupational therapists for management. Occupational therapists would then conduct a comprehensive assessment to determine the underlying deficits of these students, thus providing appropriate intervention accordingly. The findings are conducive to the development of good screening tools and the clinical decision on the best interventions to enhance children’s handwriting performance [12].

While handwriting is one of the fundamental skills for school-aged children, the quality of handwriting quality is often associated with academic success [26]. For example, incorrect or illegible handwriting can lead to a reduction of marks or grades in assignments and examinations. Slow handwriting speed also affects children’s academic performance when they cannot finish tests and examinations in time. As a consequence, children, who write poorly and fail to achieve good academic results, might be misunderstood as lazy or unmotivated in school; thus, they would suffer from low self-esteem and other psychological problems [27]. The study provides a quantitative profile of children’s handwriting performance using an objective assessment tool, SHARP, thus assisting teachers and parents to understand better the studnets’ underlying problems affecting handwriting, thus early training and remediation could be provided. Students would then feel less frustrated to cope with their handwriting deficits.

### Limitations of the study

The study has several limitations. The normative data was built based on Hong Kong students with Chinese as their mother language. Thus, a careful explanation should be applied when using this assessment tool for Chinese-speaking students in other countries. Moreover, SHARP adopted near-point copying as the assessment format. Although it is one of the most widely used handwriting assessment formats, far-point copying and dictation may also be relevant for assessing students’ handwriting performance at more advanced levels. This will require cognitive abilities such as lexical processing, phonological, morphological abilities in addition to sensorimotor skills [28]. Different formats could be adopted into further research to provide a more thorough analysis and problem detection on students’ handwriting performance. Finally, previous researchers found that hardwiring performance in pen grip, the pressure applied on the paper, and the writing within the grid might be related to proprioception/kinesthesia. Yet, it is difficult to measure pure proprioception/ kinesthesia ability [29]. Other sensorimotor skills and cognitive functions might affect children’s handwriting as well. Thus, the SHARP system can further be incorporated with other technologies, such as motion capture devices, oculomotor tracking devices, or electroencephalography to monitor brain activities during the handwriting process.

## Conclusion

This study aimed to develop an objective computer-based handwriting system, SHARP, to evaluate Chinese handwriting performance and to find out its discriminant abilities to differentiate typically developing students and those with neurodevelopmental delay, thus affecting their handwriting performance. The study also aimed to find out the relationships between handwriting performance and the sensorimotor abilities of these students. Results indicated that SHARP could be an objective tool to screen out students with handwriting problems, including the process (speed, in-air time, ground time) and the product (stroke problems, out of grid, word size etc.) It could also provide a more accurate evaluation of the students’ handwriting progression. It was also observed that handwriting performance has strong relationships with fine motor and visual perceptual skills of students. These findings echo with previous studies that handwriting difficulties are associated with the underlying deficits of students in sensorimotor skills; thus intervention could be provided targeting these deficits rather than previously by rote learning or repeated copying. Further studies could be focused more on the intervention and remediation of students.

## Data Availability

Data cannot be shared publicly because of confidentiality. Data are available from the corresponding author for researchers who meet the criteria for access to confidential data.

## Acknowledgments

The authors of this paper would like to express their sincere gratitude to schools, teachers, parents, and students that participated in this study. This study was funded by the Innovation and Technology Fund (project number: ITS/265/16FX).

